# Dose prediction for repurposing nitazoxanide in SARS-CoV-2 treatment or chemoprophylaxis

**DOI:** 10.1101/2020.05.01.20087130

**Authors:** Rajith KR Rajoli, Henry Pertinez, Usman Arshad, Helen Box, Lee Tatham, Paul Curley, Megan Neary, Joanne Sharp, Neill J Liptrott, Anthony Valentijn, Christopher David, Steve P Rannard, Ghaith Aljayyoussi, Shaun H Pennington, Andrew Hill, Marta Boffito, Stephen A Ward, Saye H Khoo, Patrick G Bray, Paul M. O’Neill, W. Dave Hong, Giancarlo Biagini, Andrew Owen

**Affiliations:** Department of Molecular and Clinical Pharmacology, Materials Innovation Factory, University of Liverpool, Liverpool, L7 3NY, UK; Department of Chemistry, University of Liverpool, Liverpool, L69 3BX, UK; Centre for Drugs and Diagnostics. Department of Tropical Disease Biology, Liverpool School of Tropical Medicine, Liverpool L3 5QA, UK; Chelsea and Westminster NHS Foundation Trust and St Stephen’s AIDS Trust 4th Floor, Chelsea and Westminster Hospital, 369 Fulham Road, London, SW10 9NH, UK; Jefferiss Research Trust Laboratories, Department of Medicine, Imperial College, London, W2 1PG, UK; Pat Bray Electrical, 260D Orrell Road, Orrell, Wigan, WN5 8QZ, UK

**Keywords:** COVID-19, SARS-CoV-2, Coronavirus, Pharmacokinetics, Lung

## Abstract

**Background:** Severe acute respiratory syndrome coronavirus 2 (SARS-CoV-2) has been declared a global pandemic by the World Health Organisation and urgent treatment and prevention strategies are needed. Many clinical trials have been initiated with existing medications, but assessments of the expected plasma and lung exposures at the selected doses have not featured in the prioritisation process. Although no antiviral data is currently available for the major phenolic circulating metabolite of nitazoxanide (known as tizoxanide), the parent ester drug has been shown to exhibit *in vitro* activity against SARS-CoV-2. Nitazoxanide is an anthelmintic drug and its metabolite tizoxanide has been described to have broad antiviral activity against influenza and other coronaviruses. The present study used physiologically-based pharmacokinetic (PBPK) modelling to inform optimal doses of nitazoxanide capable of maintaining plasma and lung tizoxanide exposures above the reported nitazoxanide 90% effective concentration (EC_90_) against SARS-CoV-2.

**Methods:** A whole-body PBPK model was constructed for oral administration of nitazoxanide and validated against available tizoxanide pharmacokinetic data for healthy individuals receiving single doses between 500 mg – 4000 mg with and without food. Additional validation against multiple-dose pharmacokinetic data when given with food was conducted. The validated model was then used to predict alternative doses expected to maintain tizoxanide plasma and lung concentrations over the reported nitazoxanide EC_90_ in >90% of the simulated population. Optimal design software PopDes was used to estimate an optimal sparse sampling strategy for future clinical trials.

**Results:** The PBPK model was validated with AAFE values between 1.01 – 1.58 and a difference less than 2-fold between observed and simulated values for all the reported clinical doses. The model predicted optimal doses of 1200 mg QID, 1600 mg TID, 2900 mg BID in the fasted state and 700 mg QID, 900 mg TID and 1400 mg BID when given with food, to provide tizoxanide plasma and lung concentrations over the reported *in vitro* EC_90_ of nitazoxanide against SARS-CoV-2. For BID regimens an optimal sparse sampling strategy of 0.25, 1, 3 and 12h post dose was estimated.

**Conclusion:** The PBPK model predicted that it was possible to achieve plasma and lung tizoxanide concentrations, using proven safe doses of nitazoxanide, that exceed the EC_90_ for SARS-CoV-2. The PBPK model describing tizoxanide plasma pharmacokinetics after oral administration of nitazoxanide was successfully validated against clinical data. This dose prediction assumes that the tizoxanide metabolite has activity against SARS-CoV-2 similar to that reported for nitazoxanide, as has been reported for other viruses. The model and the reported dosing strategies provide a rational basis for the design (optimising plasma and lung exposures) of future clinical trials of nitazoxanide in the treatment or prevention of SARS-CoV-2 infection.

## Introduction

COVID-19 is a respiratory illness caused by severe acute respiratory syndrome coronavirus 2 (SARS-CoV-2) with noticeable symptoms such as fever, dry cough, and difficulty in breathing. There are currently no effective treatment or prevention options and it has become a global health problem with more than 3.1 million cases and over 217,000 deaths as of 29^th^ April 2020 [1]. Urgent strategies are required to manage the pandemic and the repurposing of already approved medicines is likely to bring options forward more quickly than full development of potent and specific antivirals. Antiviral drugs may have application prior to or during early infection, but may be secondary to immunological interventions in later stages of severe disease [2].

Although new chemical entities (NCEs) are likely to have high potency and specificity for SARS-CoV-2, full development is time consuming, costly and attrition in drug development is high [3, 4]. Drug repurposing, where existing or investigational drugs could be used outside the scope of their original indication may present a rapid alternative to new drug development. Several examples of successful repurposing exist, including the use of the anti-angiogenic drug thalidomide for cancer and the use of mifepristone for Cushing’s disease after initially being approved for termination of early pregnancy [5, 6].

SARS-CoV-2 targets the angiotensin-converting enzyme 2 (ACE2) receptors that are present in high density on the outer surface of lung cells. Lungs are the primary site of SARS-CoV-2 replication and infection is usually initiated in the upper respiratory tract [7]. Symptoms that result in neurological, renal and hepatic dysfunction are also emerging due to the expression of ACE2 receptors in these organs [8–11]. Therefore, therapeutic concentrations of antiviral drugs are likely to be needed in the upper airways for treatment and prevention of infection, but sufficient concentrations are also likely to be required systemically for therapy to target the virus in other organs and tissues.

The scale at which antiviral activity of existing medicines are being studied for potential repurposing against SARS-CoV-2 is unprecedented [12]. The authors recently reported a holistic analysis which benchmarked reported *in vitro* activity of tested drugs against previously published pharmacokinetic exposures achievable with their licenced doses [13]. Importantly, this analysis demonstrated that the majority of drugs that have been studied for anti-SARS-CoV-2 activity are unlikely to achieve the necessary concentrations in the plasma after administration of their approved doses. While this analysis is highly influenced by the drugs selected for analysis to date and highly sensitive to the accuracy of the reported antiviral activity data, a number of candidate agents were identified with plasma exposures above the reported EC_50_/EC_90_ against SARS-CoV-2.

One such drug, nitazoxanide, is a thiazolide antiparasitic medicine used for the treatment of cryptosporidiosis and giardiasis that cause diarrhoea [14, 15], and also has reported activity against anaerobic bacteria, protozoa and other viruses [16]. Importantly, rapid deacetylation of nitazoxanide in blood means that the major systemic species of the drug *in vivo* is tizoxanide, which has not yet been studied for anti-SARS-CoV-2 activity. Notwithstanding, tizoxanide has been shown to exhibit similar in vitro inhibitory activity to nitazoxanide for rotaviruses [17], hepatitis B and C viruses [18, 19], other coronaviruses, noroviruses [20] and influenza viruses [21, 22]. As another respiratory virus, previous work on influenza may be useful to gain insight into the expected impact of nitazoxanide for SARS-CoV-2. Accordingly, the drug has been shown to selectively block the maturation of the influenza hemagglutinin glycoprotein at the post-translational stage [22, 23] and a previous phase 2b/3 trial demonstrated a reduction in symptoms and viral shedding at a dose of 600 mg BID compared to placebo in patients with uncomplicated influenza [24]. Other potential benefits of nitazoxanide in COVID-19 may derive from its impact upon the innate immune response that potentiates the production of type 1 interferons [25, 26] and bronchodilation of the airways through inhibition of TMEM16A ion channels [27]. A clinical trial (NCT04341493) started on April 6^th^ 2020 and aims to evaluate the activity of 500 mg BID nitazoxanide alone or in combination with the 4-aminoquinoline hydroxychloroquine against SARS-CoV-2 [28]. However, there are currently no data within the public domain to support this dose selection for COVID-19. Nitazoxanide is relatively safe in humans and studies showed tolerability of single oral doses up to 4 g with minimal gastrointestinal side effects. Plasma concentrations of tizoxanide have demonstrated dose proportionality, but administration in the fed state increases the plasma exposure [29]. Thus, the drug is recommended for administration with food.

Physiologically-based pharmacokinetic (PBPK) modelling is a computational tool that integrates human physiology and drug disposition kinetics using mathematical equations to inform the pharmacokinetic exposure using *in vitro* and drug physicochemical data [30]. The aim of this study was to validate a PBPK model for tizoxanide following administration of nitazoxanide. Once validated this model was first used to assess the plasma and lung exposures estimated to be achieved during a previous trial for uncomplicated influenza. Next, different nitazoxanide doses and schedules were simulated to identify those expected to provide tizoxanide plasma and lung trough concentrations (C_trough_) above the reported nitazoxanide SARS-CoV-2 EC_90_ in the majority (>90%) of patients.

## Methods

A whole-body PBPK model consisting of compartments to represent select organs and tissues was developed. Nitazoxanide physiochemical and drug-specific parameters used in the PBPK model were obtained from literature sources as outlined in Table 1. The PBPK model was assumed to be blood-flow limited, with instant and uniform distribution in each tissue or organ and no reabsorption from the large intestine. Since the data is computer generated, no ethics approval was required for this study.

**Table 1.**
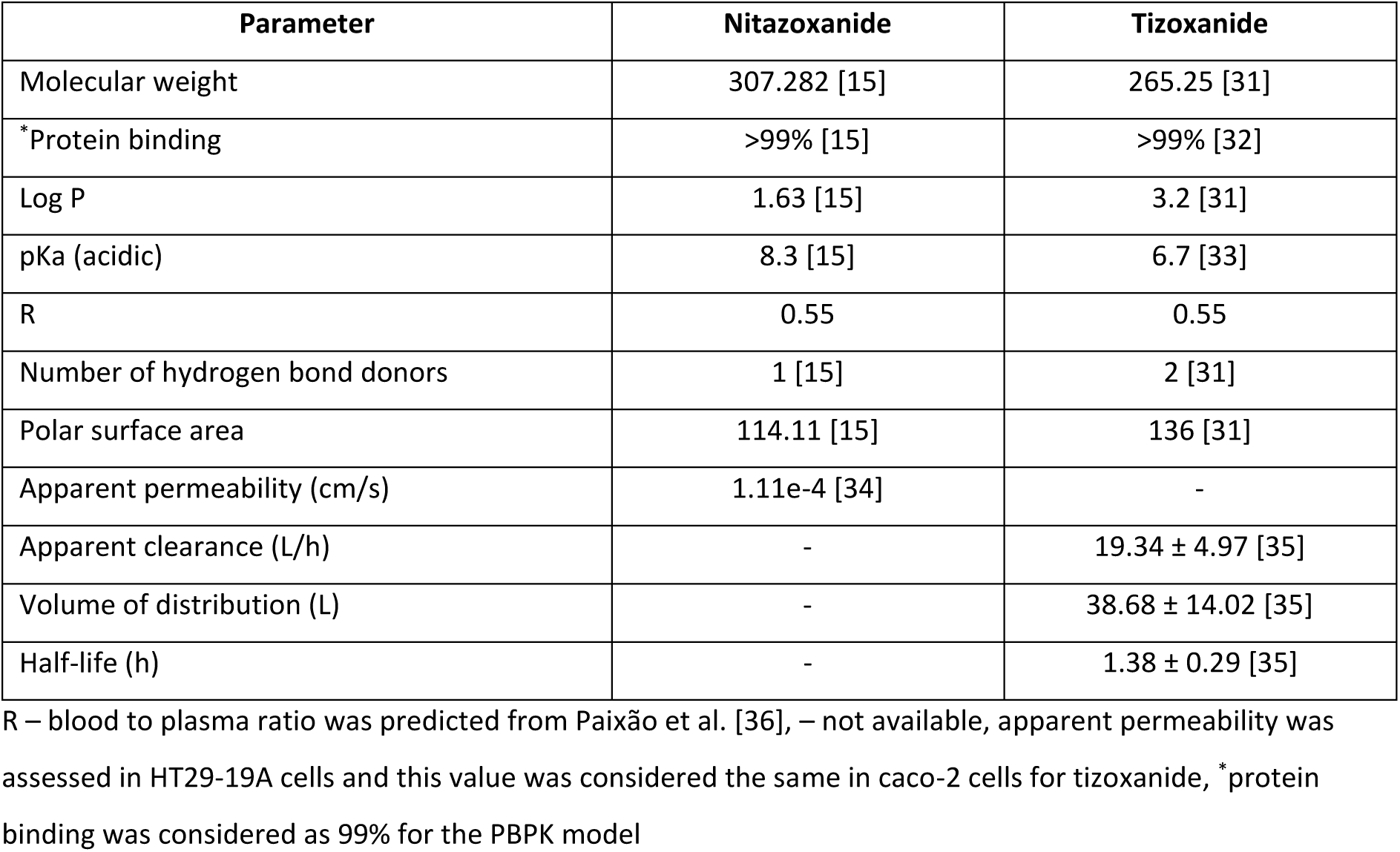
Nitazoxanide input parameters for the PBPK model

| Parameter | Nitazoxanide | Tizoxanide |
| --- | --- | --- |
| Molecular weight | 307.282 [15] | 265.25 [31] |
| *Protein binding | >99% [15] | >99% [32] |
| Log P | 1.63 [15] | 3.2 [31] |
| pKa (acidic) | 8.3 [15] | 6.7 [33] |
| R | 0.55 | 0.55 |
| Number of hydrogen bond donors | 1 [15] | 2 [31] |
| Polar surface area | 114.11 [15] | 136 [31] |
| Apparent permeability (cm/s) | 1.11e-4 [34] | - |
| Apparent clearance (L/h) | - | 19.34 ± 4.97 [35] |
| Volume of distribution (L) | - | 38.68 ± 14.02 [35] |
| Half-life (h) | - | 1.38 ± 0.29 [35] |

R – blood to plasma ratio was predicted from Paixão et al. [36], – not available, apparent permeability was assessed in HT29-19A cells and this value was considered the same in caco-2 cells for tizoxanide, \*protein binding was considered as 99% for the PBPK model

### Model development

One hundred virtual healthy adults (50% women, aged 20-60 years between 40-120 kg) were simulated. Patient demographics such as weight, BMI and height were obtained from CDC charts [37]. Organ weight/volumes and blood flow rates in humans were obtained from published literature sources [38, 39]. Transit from the stomach and small intestine was divided into seven compartments to capture effective absorption kinetics as previously described [40]. Tissue to plasma partition ratio of drug and drug disposition across various tissues and organs were described using published mathematical equations [41–43]. Effective permeability (P_eff_) in humans was scaled from apparent permeability (P_app_) in HT29-19A cells (due to lack of available data, it was assumed the same in Caco-2 cells) using the following equations to compute the rate of absorption (K_a_ in h^-1^) from the small intestine.

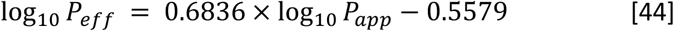

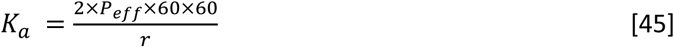

### Model validation

The PBPK model was validated against available clinical data in healthy individuals in the fed and fasted state for various single oral doses of nitazoxanide ranging from 500 mg to 4000 mg [29, 46], and for multiple dosing at 500 mg and 1000 mg BID with food. Nitazoxanide absorption was considered using the available apparent permeability data (shown in table 1) and tizoxanide was assumed to form as soon as the drug reached systemic circulation as metabolic studies have shown it takes just six minutes for complete conversion into the active circulating metabolite, with no trace of nitazoxanide detected in plasma [47]. Therefore, tizoxanide parameters were used to define drug disposition. The model was assumed to be validated if: 1) the absolute average fold error (AAFE) between the observed and the simulated plasma concentrations - time curve of tizoxanide was less than two; and 2) the simulated pharmacokinetic parameters – maximum concentration (C_max_), area under the plasma concentration-time curve (AUC) and C_trough_ (trough concentration at the end of the dosing interval) were less than two-fold from the mean observed values.

### Model simulations

The pharmacokinetics following administration of 600 mg BID as reported in a previous phase 2b/3 clinical trial of nitazoxanide in uncomplicated influenza [24] were first simulated and plotted relative to the average of previously reported influenza EC_90_s [48, 49] for strains(as shown in supplementary table 1) included in the previous trials. This was done to assess the exposure relative to *in vitro* activity for an indication where clinical benefit was already demonstrated.

For potential SARS-CoV-2 applications, several oral dosing regimens were simulated for BID, TID or QID administration in the fasted state. Antiviral activity data from Wang et al. [50] were digitised using Web Plot Digitiser^®^ software and used to calculate a nitazoxanide EC_90_ for SARS-CoV-2 of 4.64 μM (1.43 mg/L). Optimal doses were identified such that the concentration at 12 hours post-first dose (C_12_) for BID, 8 hours post-first dose (C_8_) for TID, or 6 hours post-first dose (C_6_) for QID administration were over the recalculated EC_90_ for nitazoxanide. Plasma and lung tizoxanide exposures at these doses and schedules are reported in addition to plasma-time curves. The doses were optimised using tizoxanide parameters and pharmacokinetics, however, the doses were reported for nitazoxanide.

### Optimal pharmacokinetic sampling

Clinical trials should incorporate PK sampling to confirm tizoxanide plasma exposures, and further validate the predictions from the PBPK model. Optimal sparse pharmacokinetic timepoint selection (assuming 4 blood samples per patient, and 40 patients in the study) were made on the basis of the prior fed PK data of Stockis et al. [29, 46]. Tizoxanide plasma PK data in fed patients from Stockis et al. was fitted with an empirical one compartment disposition model, with first-order absorption and absorption transit compartment, and the parameters from this fitting were used (with nominal %CV interindividual variability in the PK parameters of 30%) in the optimal design software PopDes (University of Manchester Version 4.0) to generate the suggested optimal sampling timepoints [51, 52].

## Results

### Model validation

The PBPK model validation against various fasted oral doses is shown in supplementary Figure 1 and the validation against single and multiple doses in the fed state is shown in supplementary Figure 2. The corresponding pharmacokinetic parameters (AUC, C_max_ and C_trough_) are presented in Table 2a and 2b. The AAFE values for the validated doses ranged between 1.01 – 1.55 for fasted state and between 1.1 – 1.58 for fed state indicating a close match between observed and simulated data. The ratio between the simulated and the observed pharmacokinetic parameters – AUC, C_max_ and C_trough_ was between 0.81 – 1.54 (Table 2a) for fasted state and between 0.67 – 2.15 for fed state. The PBPK model simulated tizoxanide plasma concentrations were within acceptable ranges and therefore the PBPK model was assumed to be validated.

**Table 2a.** Tizoxanide validation against observed data for various single oral doses in the fasted state

| Dose (mg) | Pharmacokinetic parameter | Observed | Simulated | Simulated/observed |
| --- | --- | --- | --- | --- |
| 500 [46] | *AUC<sub>0-12 h</sub> (µg.h/ml) | 27.02 ± 6.24 | 24.32 ± 5.07 | 0.90 |
|  | *C<sub>max</sub> (µg/ml) | 6.80 ± 1.32 | 6.47 ± 1.17 | 0.95 |
|  | †C<sub>trough</sub> (µg/ml) | 0.106 | 0.119 ± 0.185 | 1.12 |
| 1000 [29] | ^AUC<sub>0-24 h</sub> (µg.h/ml) | 50.6 (29.0 – 88.4) | 48.9 (34.3 – 63.4) | 0.93 |
|  | ^C<sub>max</sub> (µg/ml) | 12.3 (8.21 – 18.3) | 12.9 (10.2 – 12.9) | 1.05 |
|  | †C<sub>trough</sub> (µg/ml) | 0.23 | 0.21 (0.09 – 0.53) | 0.93 |
| 2000 [29] | ^AUC<sub>0-24 h</sub> (µg.h/ml) | 59.2 (36.5 – 95.9) | 68.5 (48.9 – 84.1) | 1.12 |
|  | ^C<sub>max</sub> (µg/ml) | 9.08 (7.07 – 11.7) | 9.45 (7.18 – 11.7) | 1.04 |
|  | ^†C<sub>trough</sub> (µg/ml) | 1.49 | 1.33 (0.45 – 2.21) | 0.89 |
| 3000 [29] | ^AUC<sub>0-24 h</sub> (µg.h/ml) | 52.9 (34.6 – 81.0) | 42.6 (31.7 – 53.6) | 0.81 |
|  | ^C<sub>max</sub> (µg/ml) | 7.39 (6.07 – 8.98) | 7.51 (5.68 – 9.33) | 1.02 |
|  | ^†C<sub>trough</sub> (µg/ml) | 0.6 | 0.93 (0.79 – 1.06) | 1.54 |
| 4000 [29] | ^AUC<sub>0-24 h</sub> (µg.h/ml) | 88.5 (53.5 – 146) | 76.9 (60.1 – 93.8) | 0.87 |
|  | ^C<sub>max</sub> (µg/ml) | 10.5 (8.16 – 13.5) | 9.58 (7.34 – 11.8) | 0.91 |
|  | ^†C<sub>trough</sub> (µg/ml) | 2.48 | 2.44 (2.19 – 2.67) | 0.98 |

\*C<sub>max</sub> and AUC<sub>0-12h</sub> are represented as arithmetic mean ± SD, ^C<sub>max</sub> and AUC<sub>0-24h</sub> are represented as geometric mean (mean – SD, mean + SD), †C<sub>trough</sub> is C<sub>12</sub> and has been digitised from the pharmacokinetic curve as the geometric mean is not available, arithmetic mean is shown for observed and arithmetic mean (mean - SD, mean + SD) are shown for simulated data, ^C<sub>max</sub>, AUC<sub>0-24h</sub> and ^C<sub>trough</sub> were normalised to a 1000 mg dose.

**Table 2b.** Tizoxanide validation against observed data for various single and multiple oral doses when given with food.

| Dose (mg) | Pharmacokinetic parameter | Observed | Simulated | Simulated/observed |
| --- | --- | --- | --- | --- |
| 500 (single) [53] | *AUC<sub>0-last</sub> (µg.h/ml) | 41.8 (36.1 – 48.4) | 33.2 (23.1 - 43.2) | 0.79 |
|  | C<sub>max</sub> (µg/ml) | 10.4 (8.65 – 12.6) | 8.08 (6.17 - 9.98) | 0.78 |
|  | <sup>†</sup>C<sub>trough</sub> (µg/ml) | 0.178 | 0.28 (0 - 0.5) | 1.56 |
| 500 BID<br>(multiple) [53] | *AUC<sub>0-12 h</sub> (µg.h/ml) | 48.7 (36.0 – 65.9) | 32.8 (19.5 - 46.1) | 0.67 |
|  | C<sub>max</sub> (µg/ml) | 9.05 (7.13 – 11.5) | 7.66 (5.89 - 9.44) | 0.85 |
|  | <sup>†</sup>C<sub>trough</sub> (µg/ml) | 0.310 | 0.35 (0 - 0.89) | 1.11 |
| 1000 (single)<br>[53] | *AUC<sub>0-last</sub> (µg.h/ml) | 85.9 (68.8 – 107) | 91.3 (69 - 113.5) | 1.06 |
|  | C<sub>max</sub> (µg/ml) | 14.4 (10.8 – 19.2) | 17.8 (14.3 - 21.3) | 1.23 |
|  | <sup>†</sup>C<sub>trough</sub> (µg/ml) | 0.786 | 1.43 (0.2 - 2.22) | 1.82 |
| 1000 BID<br>(multiple) [53] | *AUC<sub>0-12 h</sub> (µg.h/ml) | 144 (105 – 198) | 90.9 (69 - 112.8) | 0.63 |
|  | C<sub>max</sub> (µg/ml) | 24.2 (20.3 – 28.7) | 18.2 (14.4 - 21.9) | 0.75 |
|  | <sup>†</sup>C<sub>trough</sub> (µg/ml) | 1.68 | 1.38 (0.33 - 2.07) | 0.82 |
| 2000 (single)<br>[29] | <sup>^</sup>AUC<sub>0-24 h</sub> (µg.h/ml) | 110 (88.0 – 139) | 109 (74.8 - 142) | 0.99 |
|  | <sup>^</sup>C<sub>max</sub> (µg/ml) | 15.8 (13.0 – 19.2) | 14.3 (11.2 - 17.4) | 0.91 |
|  | <sup>^†</sup>C<sub>trough</sub> (µg/ml) | 1.52 | 3.27 (1.73 - 4.46) | 2.15 |
| 3000 (single)<br>[29] | <sup>^</sup>AUC<sub>0-24 h</sub> (µg.h/ml) | 95.3 (60.0 – 152) | 90 (60.9 - 119) | 0.94 |
|  | <sup>^</sup>C<sub>max</sub> (µg/ml) | 10.0 (7.40 – 13.5) | 10.4 (8.09 - 12.7) | 1.04 |
|  | <sup>^†</sup>C<sub>trough</sub> (µg/ml) | 2.35 | 2.64 (1.44 - 3.56) | 1.12 |
| 4000 (single)<br>[29] | $\wedge$AUC<sub>0-24 h</sub> (μg.h/ml) | 192 (99.5 – 370) | 161 (125 - 196) | 0.84 |
| | $\wedge$C<sub>max</sub> (μg/ml) | 17.5 (11.5 – 26.5) | 14.2 (11.2 - 17.2) | 0.81 |
| | $\wedge^+$C<sub>trough</sub> (μg/ml) | 6.55 | 6.52 (5.04 - 7.84) | 0.99 |

C<sub>max</sub> and AUC are represented as geometric mean (mean – SD, mean + SD) and $\wedge$ C<sub>max</sub> and AUC<sub>0-24h</sub> were normalised to a 1000 mg dose, $\wedge$ C<sub>max</sub> and AUC<sub>0-24h</sub> were normalised to a 1000 mg dose, \*AUC is represented as AUC<sub>0-∞</sub> after the first dose for single and AUC<sub>0-12</sub> on day 7, $\wedge^+$ C<sub>trough</sub> is C<sub>12</sub> has been digitised from the pharmacokinetic curve as the geometric mean is not available, arithmetic mean is shown for observed and arithmetic mean (mean - SD, mean + SD) are shown for simulated data.

### Model simulations

Figure 1a and 1b show the simulated plasma and lung exposures relative to the average influenza EC_90_ after administration of 600 mg BID dose of nitazoxanide with food as reported in the previous phase 2b/3 trial in uncomplicated influenza [24]. These simulations indicate that all patients were predicted to have plasma and lung tizoxanide C_trough_ (C_12_) concentrations below the average EC_90_ (8.4 mg/L, supplementary table 1) [49], but that 71% and 14% were predicted to have plasma and lung C_max_ concentrations respectively above the average EC_90_ for influenza, respectively.

**Figure 1.**
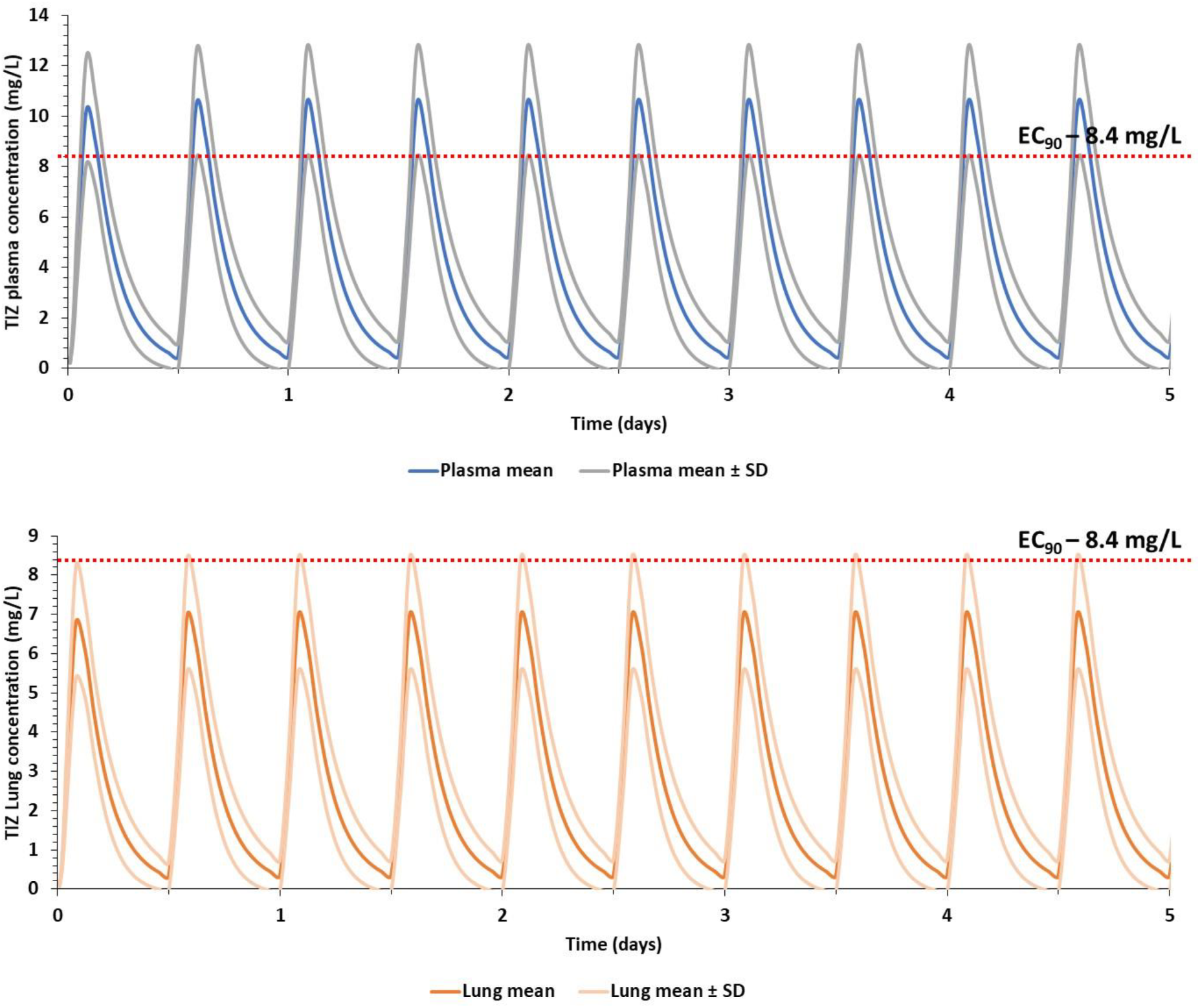
Stimulated plasma (a) and lung (b) concentration for nitazoxanide 600 mg BID for 5 days with food relative to the average reported tizoxanide EC_90_ value for influenza strains (supplementary table 1) similar to those in a previous phase IIb/III trial in uncomplicated influenza [24].

Figure 2a and 2b shows the prediction of trough concentrations in plasma and lung for the different simulated doses and schedules in healthy individuals for fasted and fed states, respectively. Doses and schedules estimated to provide plasma C_trough_ concentrations over 1.43 mg/L for at least 50% of the simulated population were identified. However, lower doses in each schedule (i.e. 800 mg QID, 1300 mg TID and 1800 mg BID in fasted state and 500 mg QID, 700 mg TID, 1100 mg BID in fed state) were predicted to result in >40% of the simulated population having lung C_trough_ below the SARS-CoV-2 EC_90_. Optimal doses for SARS-CoV-2 in the fasted state were predicted to be 1200 mg QID, 1600 mg TID, 2900 mg BID and in the fed state were 700 mg QID, 900 mg TID and 1400 mg BID. Figure 3 shows the plasma and lung concentrations for the optimal doses and schedules in fed state and supplementary figure 3 shows the plasma concentration – time profile of optimal doses in fasted state. Tizoxanide concentrations in lung and plasma were predicted to reach steady state in <48 hours, both in the fasted and fed state.

**Figure 2a.**
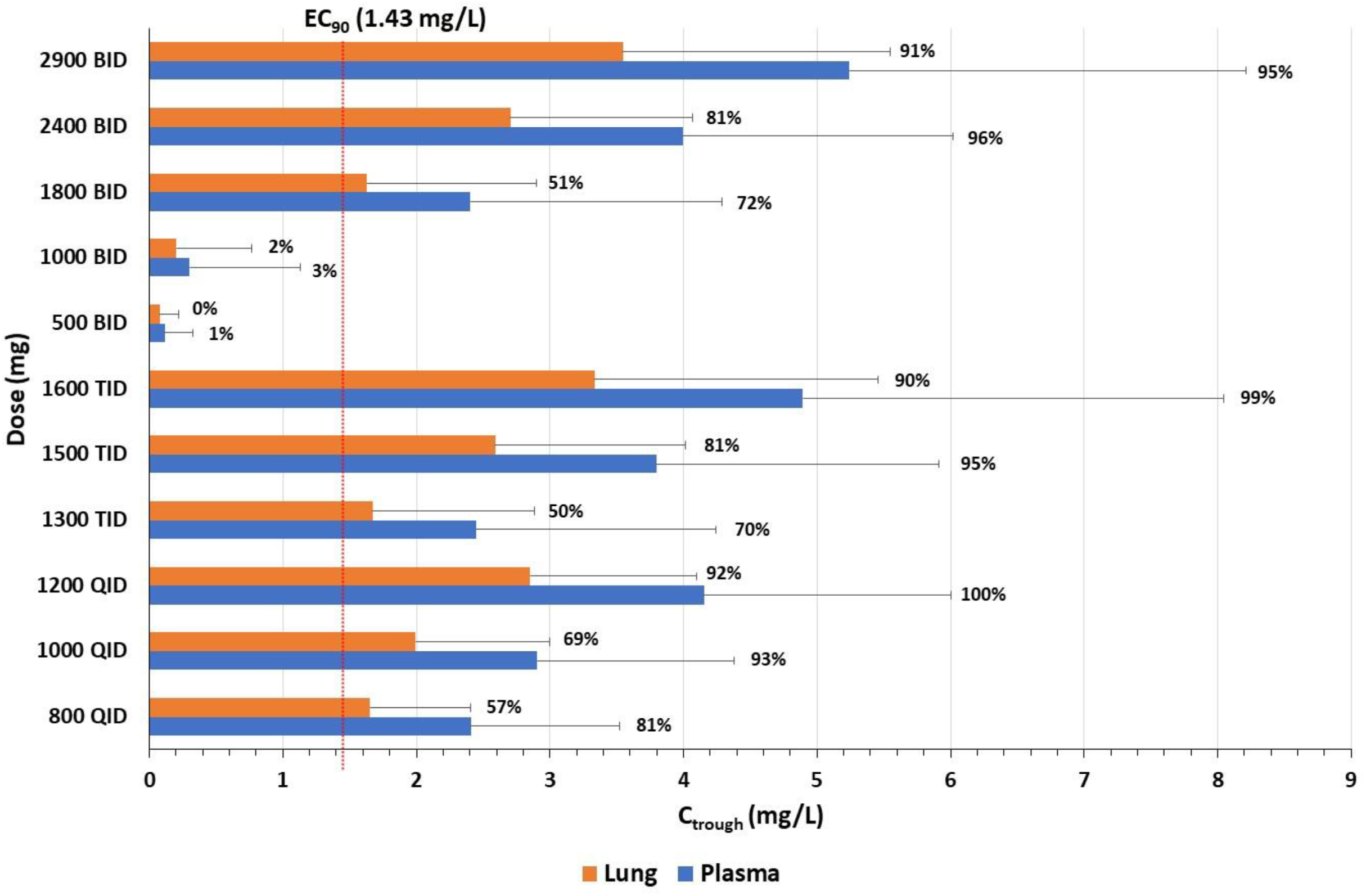
Predicted tizoxanide C_trough_ (BID - 12 h, TID - 8 h, QID - 6 h) for various dosing regimens of nitazoxanide in fasted state at the end of the first dose. Data is presented as mean and error bars represent standard deviation. The percentages adjacent to the bar chart indicate the percentage of simulated population over EC_90_ of nitazoxanide for SARS-CoV-2.

**Figure 2b.**
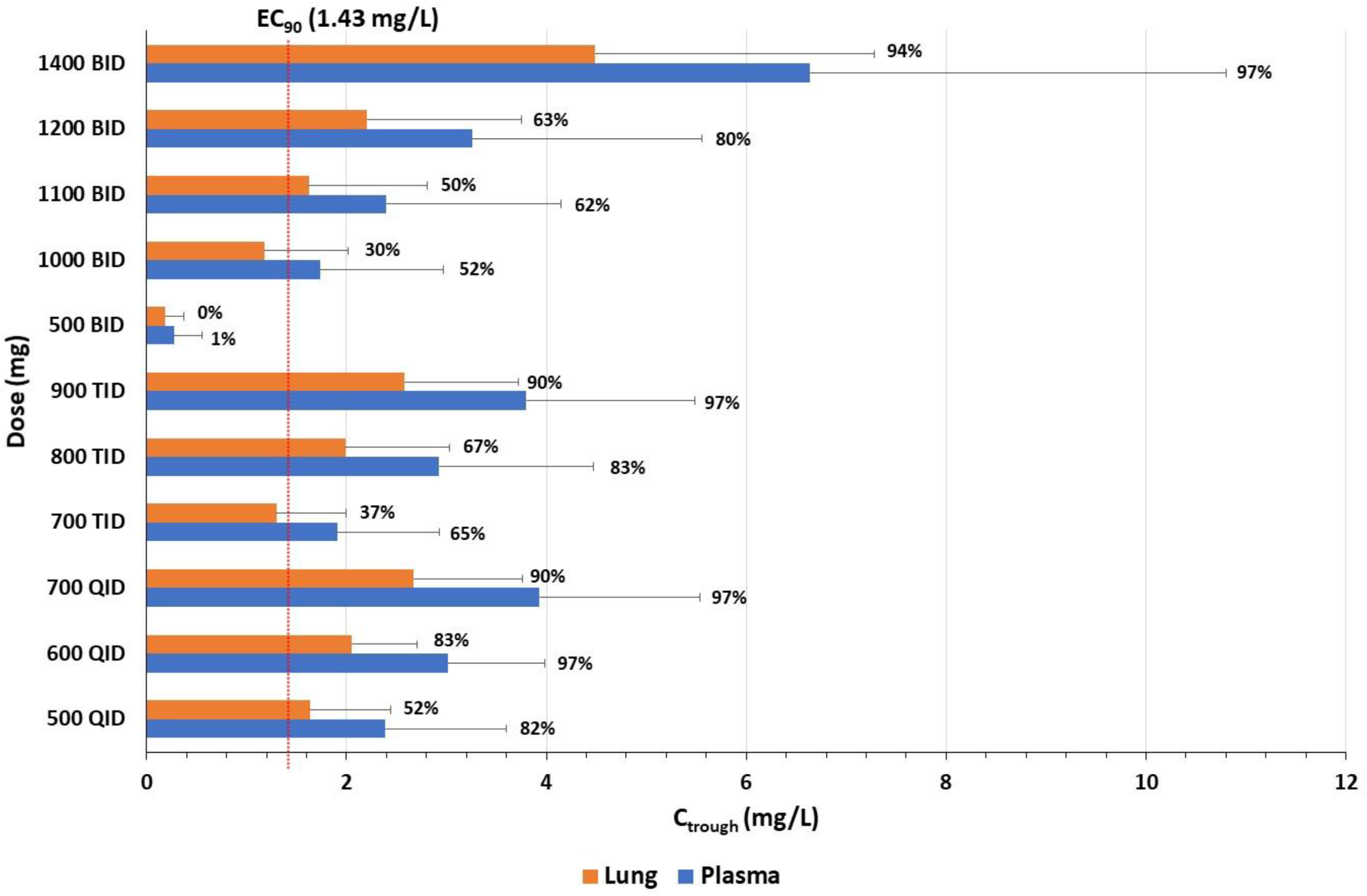
Predicted tizoxanide C_trough_ (BID – 12 h, TID – 8 h, QID – 6 h) for various dosing regimens of nitazoxanide in fed state at the end of the first dose. Data is presented as mean and error bars represent standard deviation. The percentages adjacent to the bar chart indicate the percentage of simulated population over EC_90_ of nitazoxanide for SARS-CoV-2.

**Figure 3.**
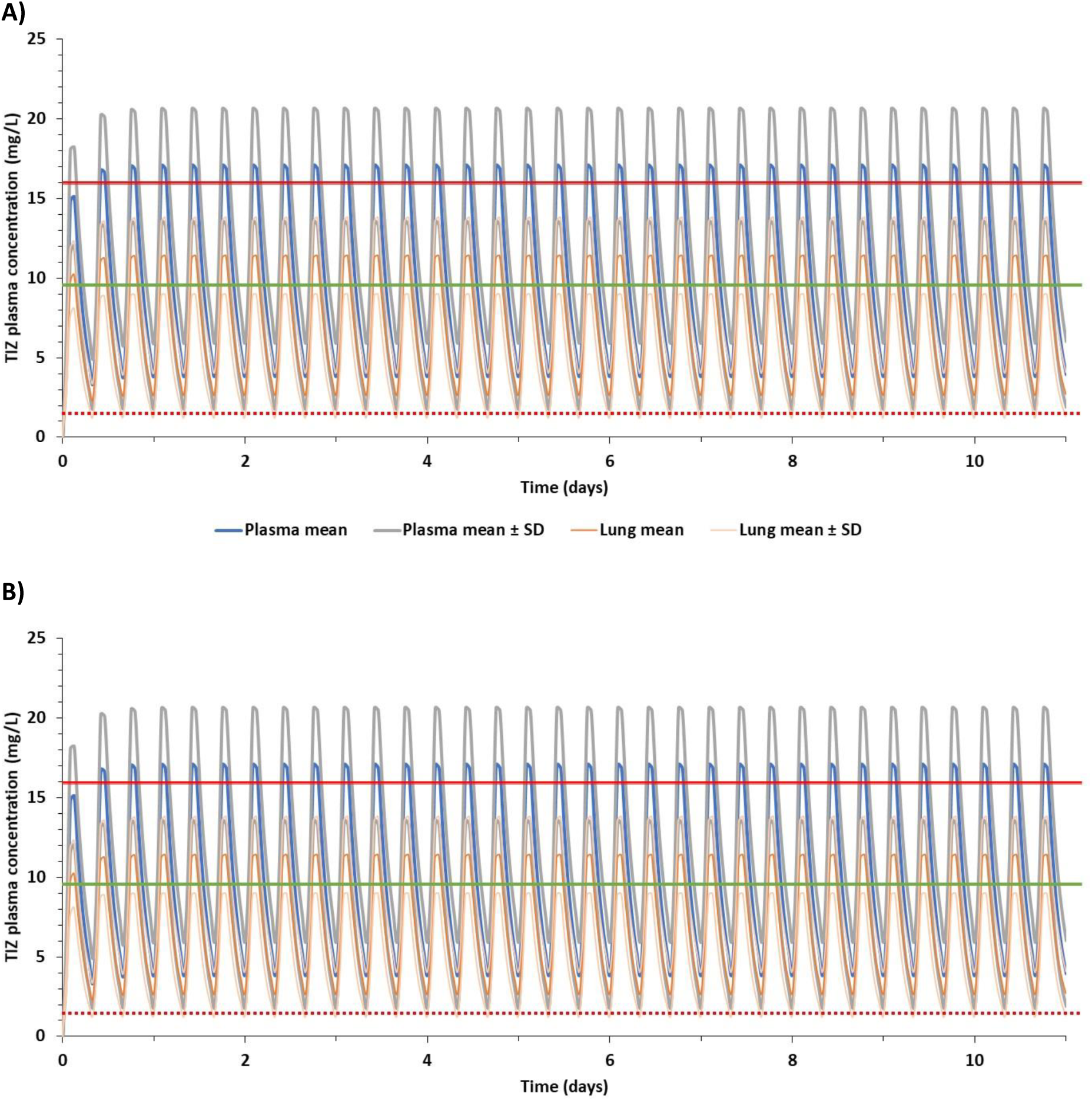
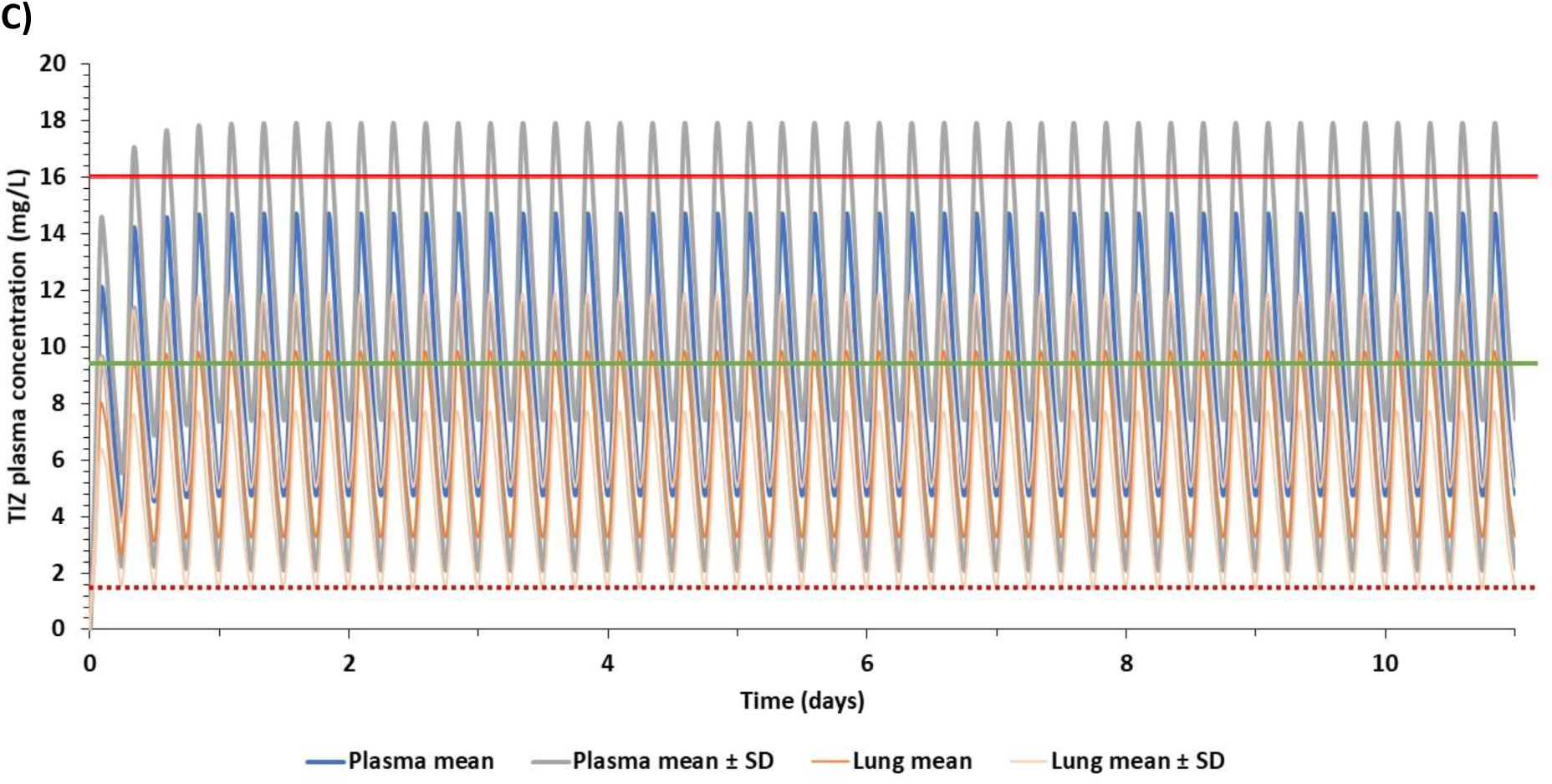
Predicted plasma and lung concentrations for optimal doses during the fed state at different regimens reaching steady state – A) 1400 mg BID, B) 900 mg TID and C) 700 mg QID. TIZ – tizoxanide, SD - standard deviation, solid red line indicates clinical C_max_ of 1 g single dose at fed state [29], solid green line represents clinical C_max_ of 500 mg single dose [53] at fed state and the dotted red line represents the EC_90_ of nitazoxanide for SARS-CoV-2 [50].

### Optimal sparse sampling design

Results from PopDes optimal design procedure indicate pharmacokinetic sampling timepoints at 0.25, 1, 3 and 12h post dose for BID regimens, and 0.25, 1, 2 and 8h post dose for TID regimens.

## Discussion

Treatment of SARS-CoV-2 has become a major global healthcare challenge with no well-defined therapeutic agents to either treat or prevent the spread of the infection. Short-term treatment options are urgently required but many ongoing trials are not based upon a rational selection of candidates in the context of safe achievable drug exposures. In the absence of a vaccine, there is also an urgent need for chemo preventative strategies to protect those at high risk such as healthcare staff, key workers and household contacts who are more vulnerable to infection. Nitazoxanide has emerged as a potential candidate for repurposing for COVID-19. The PBPK model presented herein was validated with an acceptable variation in AAFE and simulated/observed ratio (close to 1), which provides confidence in the presented predictions. The present study aimed to define the optimal doses and schedules for maintaining tizoxanide plasma and lung concentrations above the reported nitazoxanide EC_90_ for the duration of the dosing interval.

Nitazoxanide was assessed in a double-blind, randomised, placebo-controlled, phase 2b/3 trial (NCT01227421) of uncomplicated influenza in 74 primary care clinics in the USA between 27 Dec 2010 and 30 April 2011 [54]. The median duration of symptoms for patients receiving placebo was 117 h compared with 96 h in patients receiving 600 mg BID nitazoxanide with food. Importantly, virus titre in nasopharyngeal swabs in 39 patients receiving nitazoxanide 600 mg BID were also lower than in 41 patients receiving placebo. The average of reported tizoxanide EC_90_s for influenza A and B [55] was calculated to be 8.4 mg/L, which is higher than the one reported EC_90_ for nitazoxanide against SARS-CoV-2 [50]. The PBPK model was used to simulate plasma and lung exposures after administration of 600 mg BID for 5 days, and while only plasma C_max_ exceeded the average influenza EC_90_ in the majority of patients, the C_trough_ values did not. The modelling data suggest that the moderate effects of nitazoxanide seen in influenza could be a function of under dosing. Taken collectively, these data are encouraging for the application of nitazoxanide in COVID-19, assuming that tizoxanide displays anti-SARS-CoV-2 activity comparable to that reported for nitazoxanide. Moreover, these simulations indicate that higher doses may be optimal for maximal suppression of pulmonary viruses.

In some cases, food intake may be difficult in patients with COVID-19 so drugs that can be given without regard for food may be preferred. However, the presented predictions indicate that optimal plasma and lung exposures would require 1200 mg QID, 1600 mg TID or 2900 mg BID in the fasted state. Conversely, the PBPK models predict that doses of 700 mg QID, 900 mg TID or 1400 mg BID with food provide tizoxanide concentrations in plasma and lung above the EC_90_ value for nitazoxanide for the entire dosing interval in at least 90% of the simulated population. Single doses up to 4000 mg have been administered to humans previously [29] but the drug is usually administered at 500 mg BID.. The PBPK model simulations indicate a high BID dose of 1400 mg (fed) and caution may be needed for gastrointestinal intolerance at this dose. The simulations indicate that lower TID and QID dosing regimens may also warrant investigation, and 900 mg TID as well as 700 mg QID (both with food) regimen are also predicted to provide optimal exposures for efficacy. Importantly, the overall daily dose was estimated to be comparable between the different optimal schedules and it is unclear whether splitting the dose will provide gastrointestinal benefits. For prevention application where individuals will need to adhere to regimens for longer durations, minimising the frequency of dosing is likely to provide adherence benefits. However, for short term application in therapy, more frequent dosing may be more acceptable to minimise gastrointestinal intolerance.

Nitazoxanide mechanism of action for SARS-CoV-2 is currently unknown. However, for influenza it has been reported to involve interference with N-glycosylation of hemagglutinin [22, 55, 56]. Since the SARS-CoV-2 spike protein is also heavily glycosylated [57] with similar cellular targets in the upper respiratory tract, a similar mechanism of action may be expected [7, 58]. An ongoing trial in Mexico, is being conducted with 500 mg BID nitazoxanide with food [28] but these doses may not be completely optimal for virus suppression across the entire dosing interval.

This analysis provides a rational dose optimisation for nitazoxanide for treatment and prevention of COVID-19. However, there are some important limitations that must be considered. PBPK models can be useful in dose prediction but the quality of predictions is only as good as the quality of the available data on which they are based. Furthermore, the mechanism of action for nitazoxanide for other viruses has also been postulated to involve an indirect mechanism through amplification of the host innate immune response [59], and this would not have been captured in the in vitro antiviral activity that informed the target concentrations for this dose prediction. The simulated population used in this modelling consisted of healthy individuals up to 60 years old, but many patients requiring therapy may be older and have underlying comorbidities. To best knowledge, the impact of renal and hepatic impairment on pharmacokinetics of this drug have not been assessed and may impact the pharmacokinetics. Although the current PBPK model is validated against various single doses in the fasted state and few multiple doses when given with food, the model may predict with less accuracy for multiple doses due to the unavailability of clinical data for multiple dosing over 1000 mg. The presented models were validated using BID doses only, and confidence in the predictions for TID and QID doses may be lower. The clinical studies used for model validation were performed in a limited number of patients [29] and thus may underrepresent real inter-subject variability. Also, the disposition parameters (apparent clearance and rate of absorption) obtained for the PBPK model were from a fasted study of 500 mg BID, and the parameters were adjusted to validate the tizoxanide model in the fed state, which may limit confidence in the model at higher doses. Only one manuscript has described the *in vitro* activity of nitazoxanide against SARS-CoV-2 [50] and no data are available for tizoxanide. Reported in vitro data may vary across laboratories and due to this the predicted optimal doses may change. However, the reported comparable activity of nitazoxanide and tizoxanide against a variety of other viruses (including other coronaviruses) does strengthen the rational for investigating this drug for COVID-19 [17–19, 21, 22]. Finally, none of the reported EC_90_ values for influenza or SARS-CoV-2 were protein binding-adjusted [50] and tizoxanide is known to be highly protein bound (>99%) in plasma [32]. Therefore, while the protein binding was used to estimate drug penetration into the lung, data were not available to correct the *in vitro* activity.

In summary, the developed PBPK model of nitazoxanide was successfully validated against clinical data and based on currently available data, optimal doses for COVID-19 were estimated to be 700 mg QID, 900 mg TID or 1400 mg BID with food. Should nitazoxanide be progressed into clinical evaluation for treatment and prevention of COVID-19, it will be important to further evaluate the pharmacokinetics in these population groups. In treatment trials particularly, intensive pharmacokinetic sampling may be challenging. Therefore, an optimal sparse sampling strategy for BID, TID and QID dosing is also presented.

## Data Availability

The data referred to in the manuscript has been cited accordingly as recorded in the references section of the manuscript

## Study Highlights

### What is the current knowledge on the topic?

COVID-19, an acute respiratory infection caused by SARS-CoV-2, has been declared as a pandemic by the World Health Organisation. Several repurposed drugs are being evaluated but currently there are no robustly validated treatment or preventative medicines or regimens. Nitazoxanide has shown in vitro activity against SARS-CoV-2, influenza and several other animal and human RNA viruses.

### What question did this study address?

The manuscript describes the pharmacokinetics of tizoxanide, the active metabolite of nitazoxanide using a physiologically-based pharmacokinetic (PBPK) model. The validated PBPK model was used to estimate the optimal doses required for SARS-CoV-2 treatment or prevention such that >90% of the simulated population would have tizoxanide concentrations in the plasma and lung above the reported 90% effective concentration (EC_90_) value of nitazoxanide for the entire dosing interval.

### What does this study add to our knowledge?

There are no reported studies that identify treatment regimens of nitazoxanide for SARS-CoV-2 treatment or prevention. The current study provides evidence-based alternative dosing regimens of nitazoxanide for SARS-CoV-2 treatment and prevention that exceeds antiviral EC_90_s in key tissues and organs for the duration of the dosing interval.

### How might this change clinical pharmacology or translational science?

Efficacy of nitazoxanide has been demonstrated for uncomplicated influenza and the presented predictions may help inform nitazoxanide dose selection for COVID19 clinical trials.

**Supplementary Table 1.**

| Drug | EC<sub>50</sub> (μM) | EC<sub>50</sub> (ng/ml) | EC<sub>90</sub> (μM) | EC<sub>90</sub> (ng/ml) | Influenza virus strain | Reference |
| --- | --- | --- | --- | --- | --- | --- |
| Nitazoxanide | 3.2 | 983 | 26 | 7989 | A/Puerto Rico/8/1934 (H1N1) | [48] |
|  | 1.6 | 491 | 16.3 | 5008 | A/WSN/1933 (H1N1) |  |
|  | 3.2 | 983 | 20.8 | 6391 | A/California/7/2009 (H1N1pdm09) |  |
|  | 1.9 | 583 | 21.1 | 6483 | Oseltamivir-resistant<br>A/Parma/24/2009 (H1N1) |  |
|  | 3.2 | 983 | 26.8 | 8235 | A/Goose/Italy/296246/2003(H1N1) |  |
|  | 1 | 307 | 13 | 3994 | A/Parma/06/2007(H3N2) |  |
| **Average** | **2.4** | **722** | **20.7** | **6350** | **(for nitazoxanide)** |  |
| Tizoxanide | 3.8 | 1000 | 33.9 | 9000 | H1N1-PR8 | <u>[49]</u> |
|  | 1.9 | 500 | 22.6 | 6000 | H1N1-WSN |  |
|  | 1.5 | 400 | 13.5 | 3600 | H1N1-OST-R |  |
|  | 5.7 | 1500 | 26.4 | 7000 | H1N1-A/GO |  |
|  | 3.8 | 1000 | 75.4 | 20000 | H3N2-A/FI |  |
|  | 1.1 | 300 | 26.3 | 7000 | H3N2-AMD-R |  |
|  | 3.4 | 900 | 22.6 | 6000 | FLU-B |  |
| **Average** | **3.0** | **800** | **31.6** | **8371** | **(for tizoxanide)** |  |

**Supplementary Figure 1.**
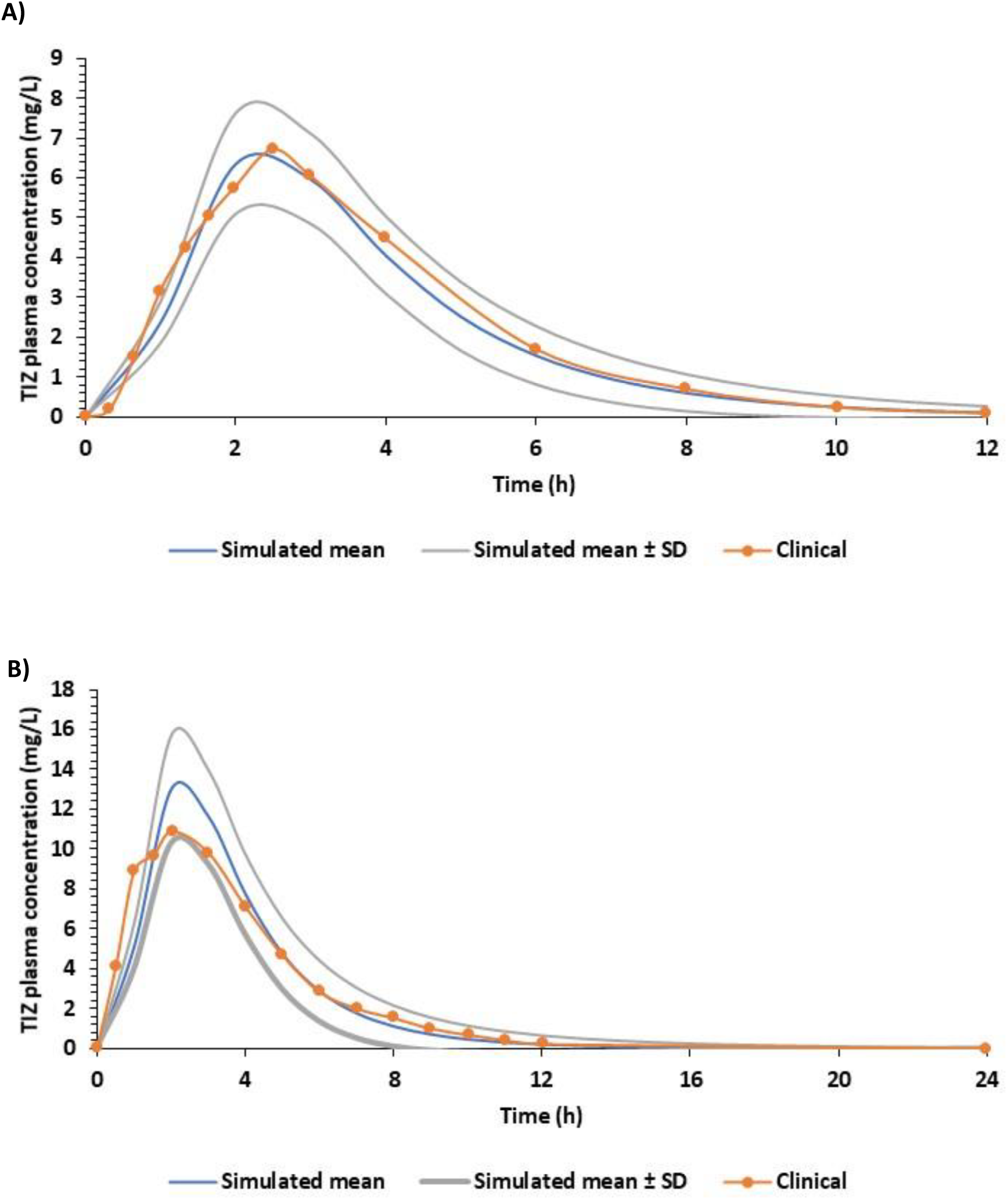
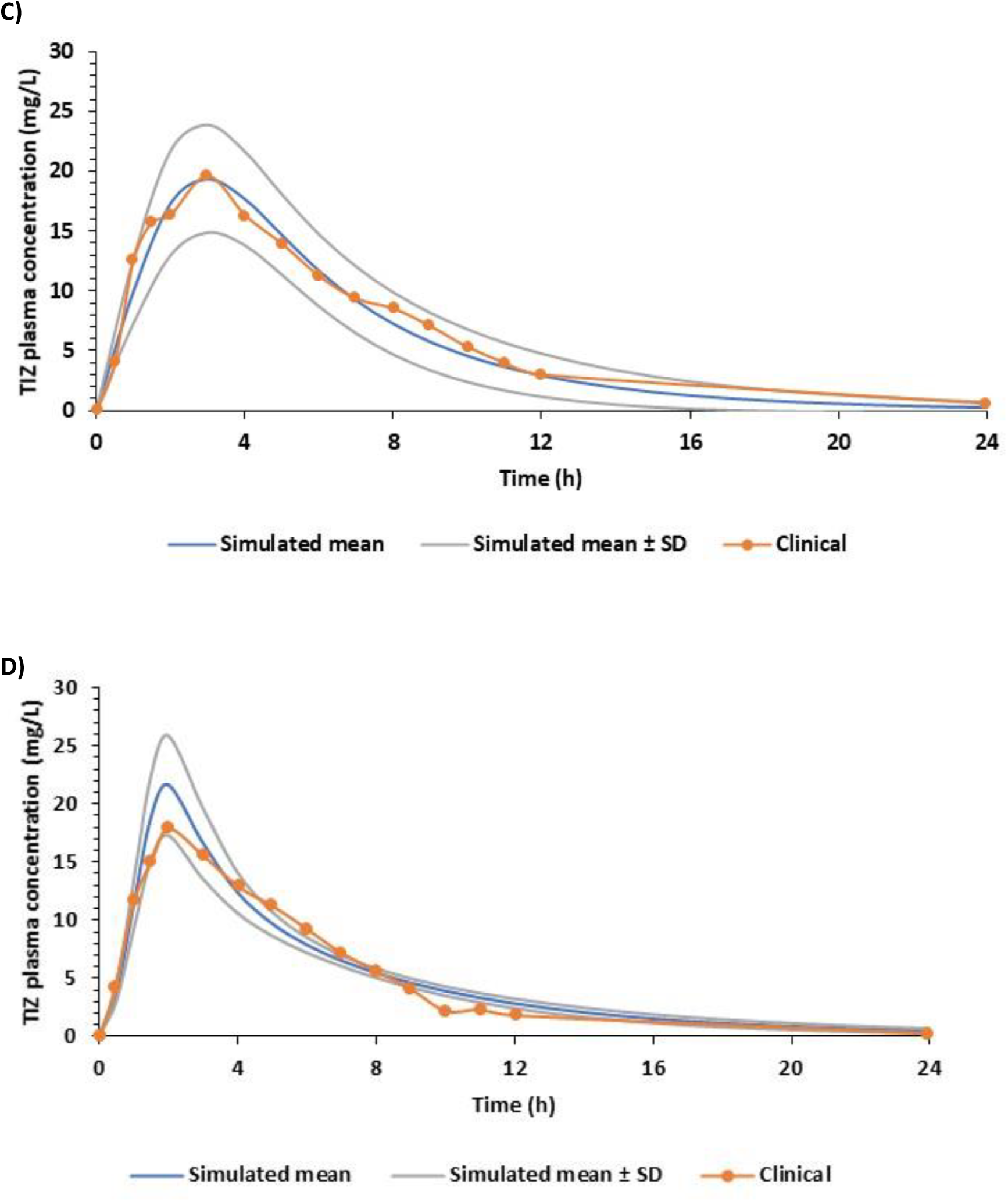
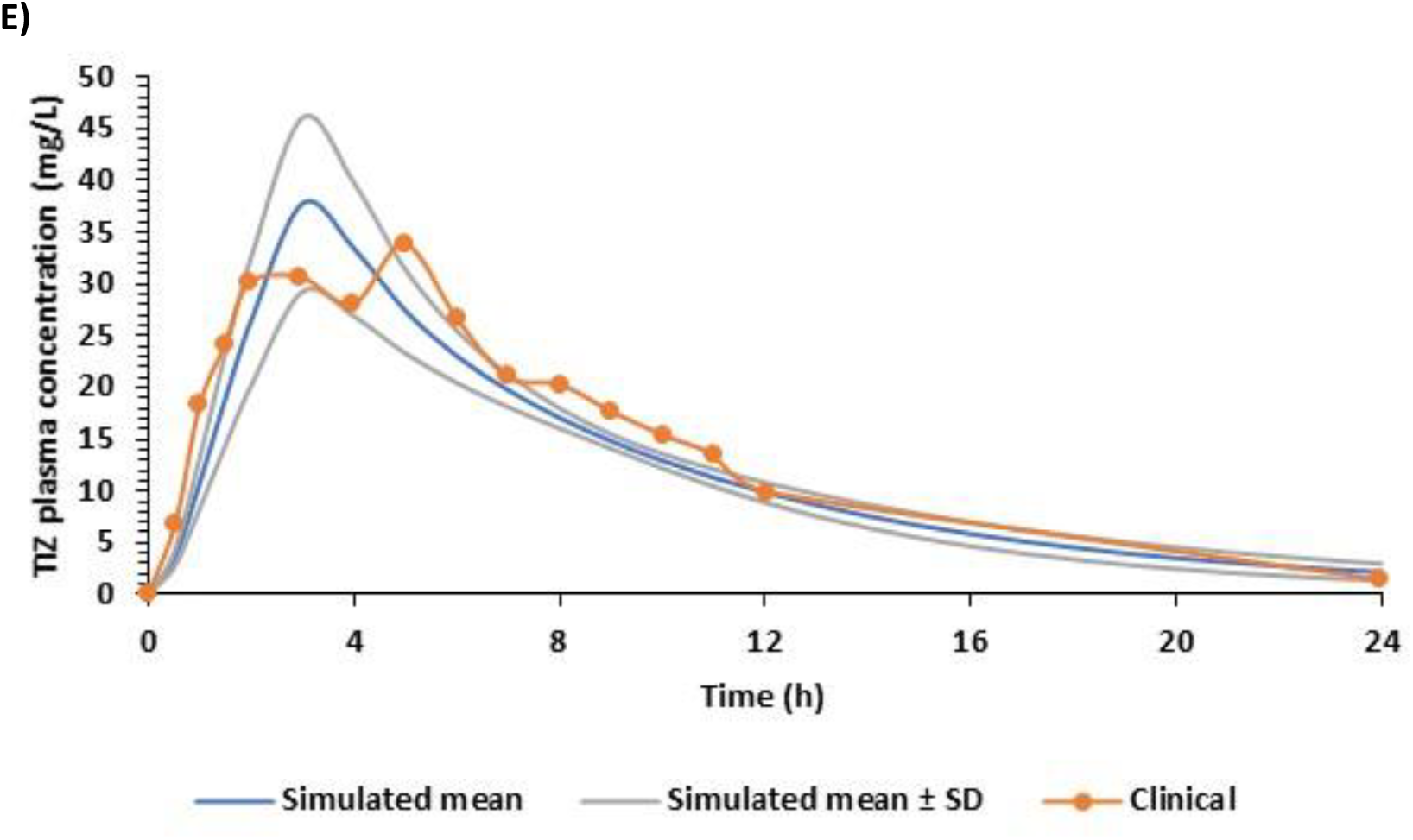
Comparison of simulated and observed plasma concentration - time curve of tizoxanide (TIZ) at fasted state. A) 500 mg, B) 1000 mg, C) 2000 mg, D) 3000 mg and E) 4000 mg

**Supplementary Figure 2.**
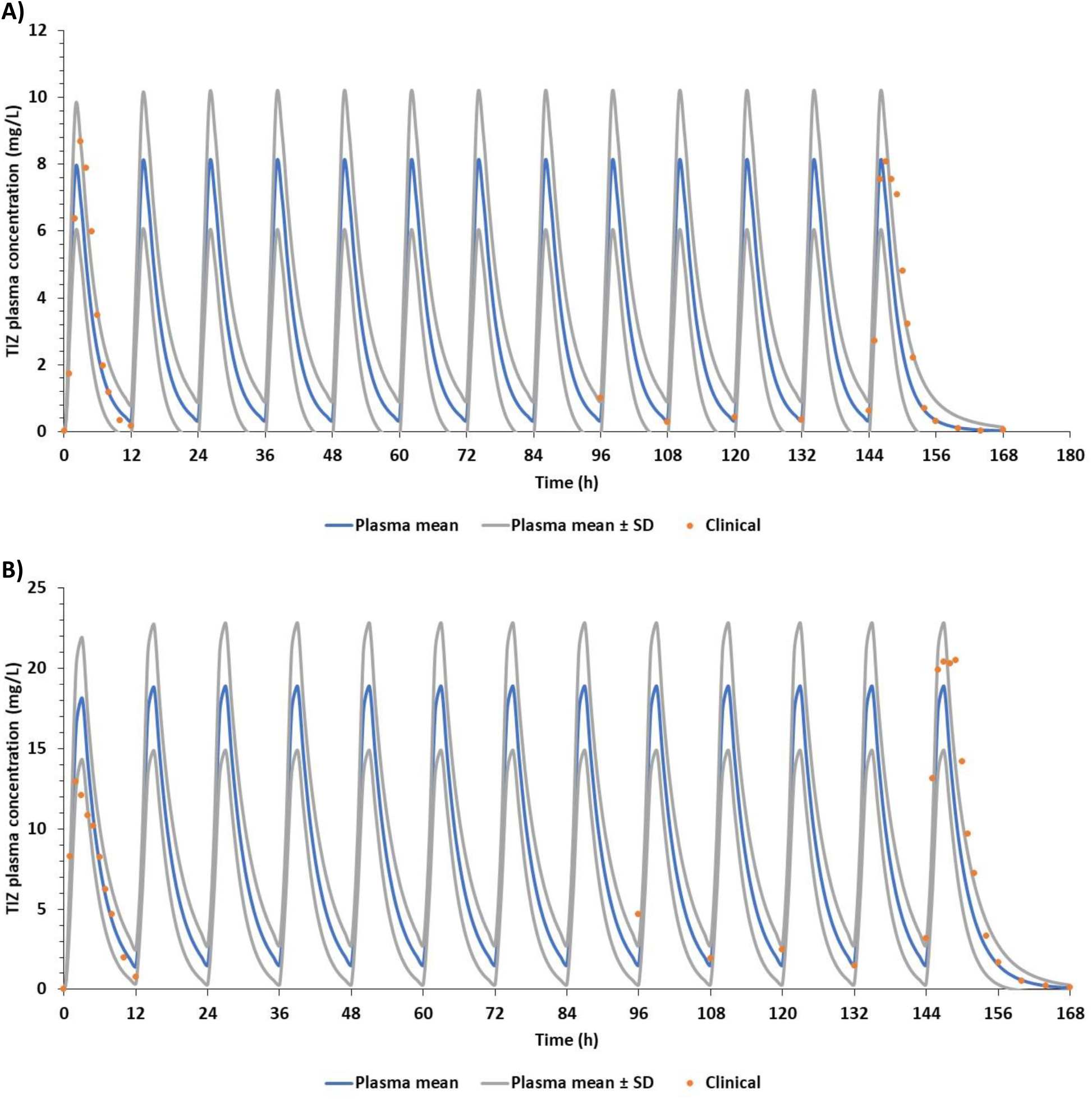
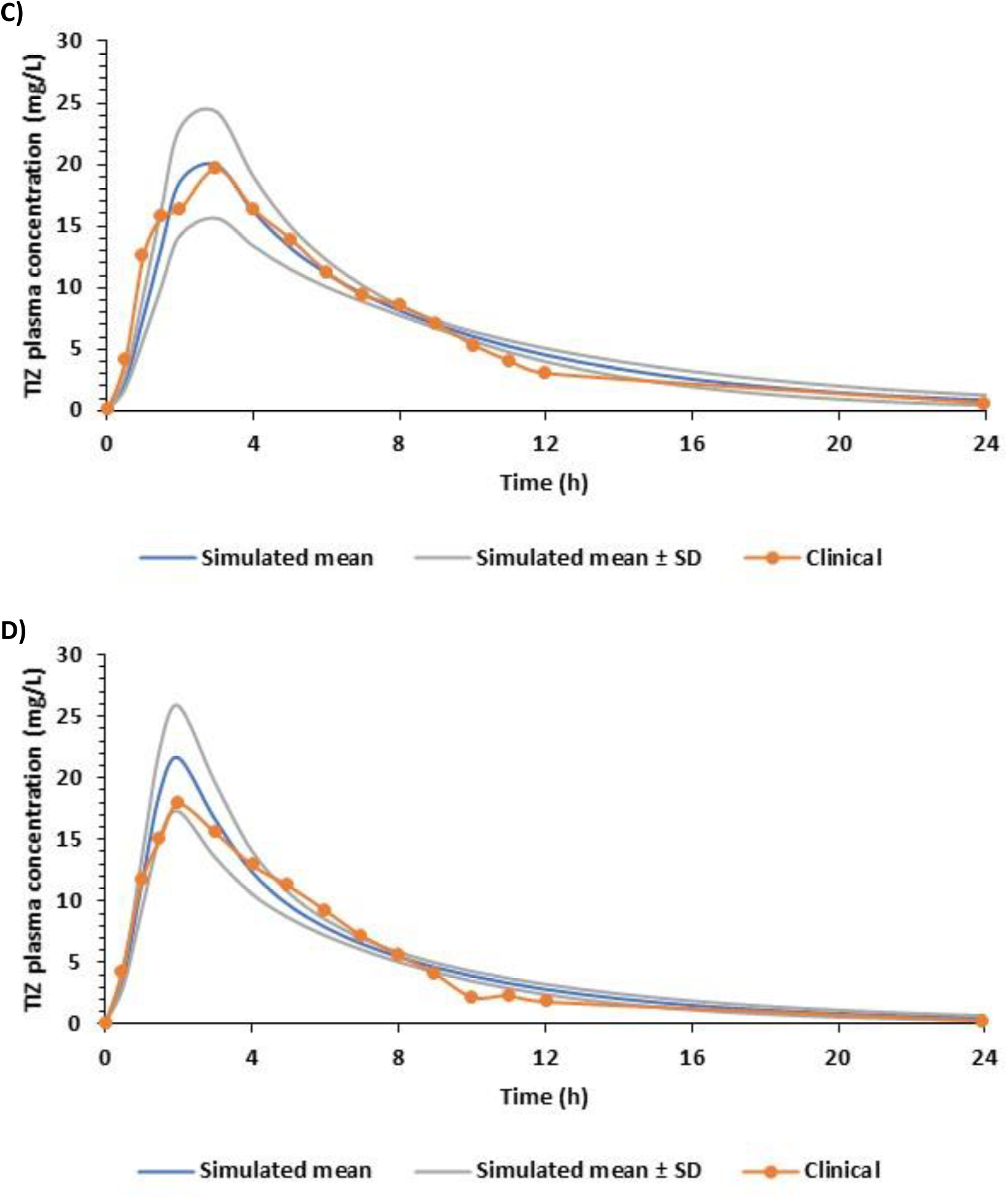
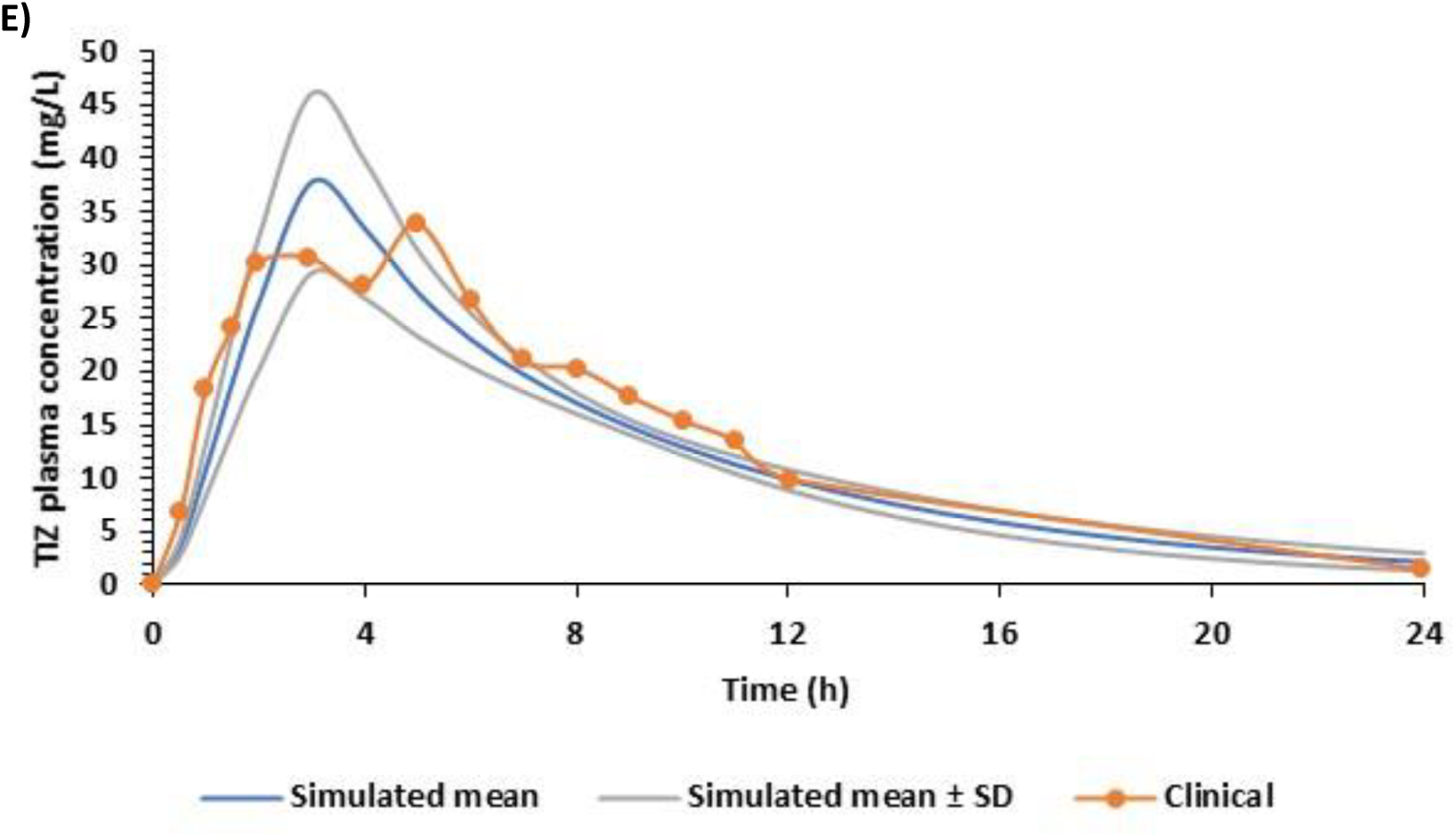
Comparison of simulated and observed plasma concentration - time curve of tizoxanide (TIZ) for single and multiple dosing regimen (where available) at fed state. A) 500 mg, B) 1000 mg, C) 2000 mg, D) 3000 mg and E) 4000 mg572

**Supplementary Figure 3.**
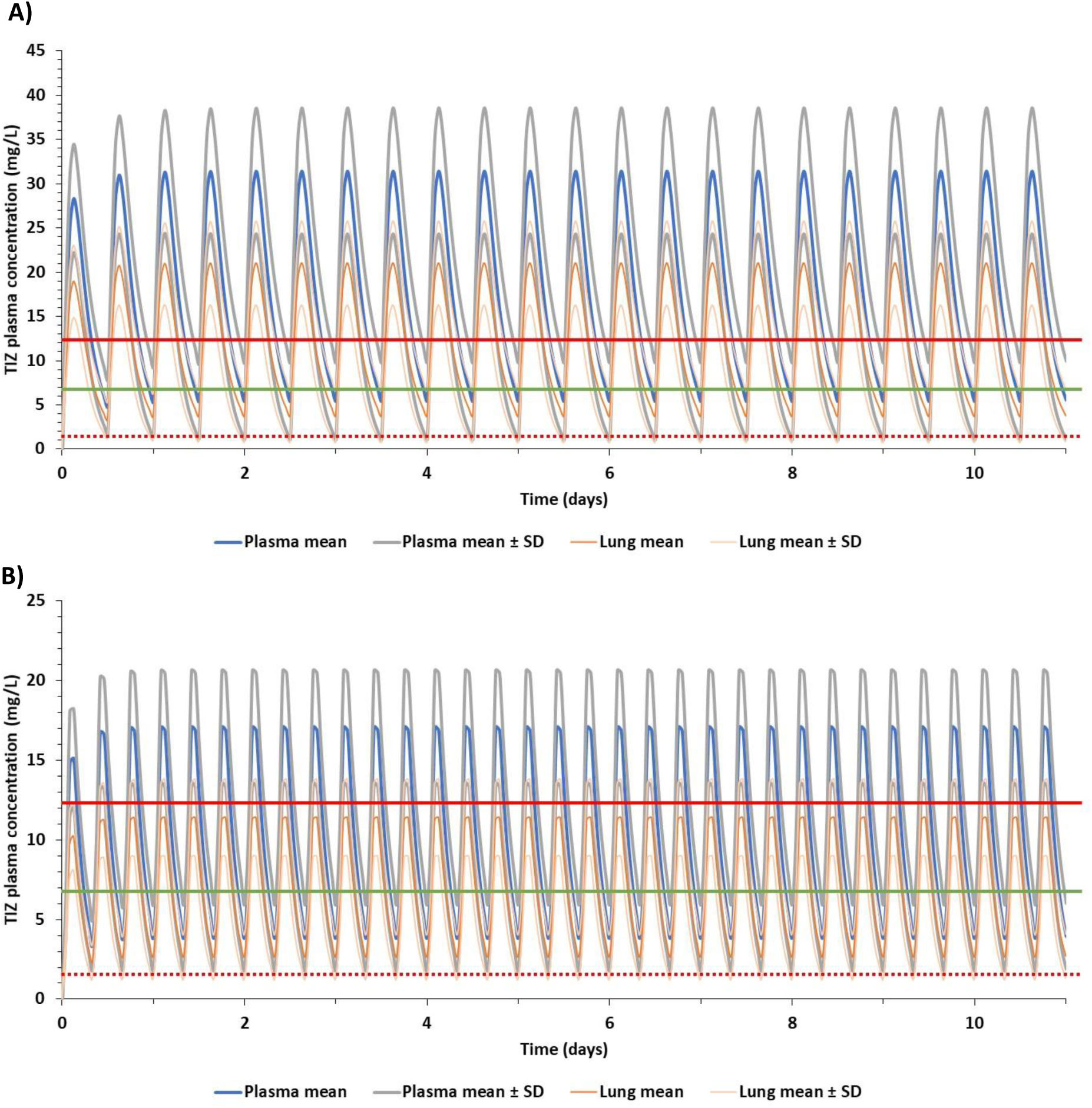
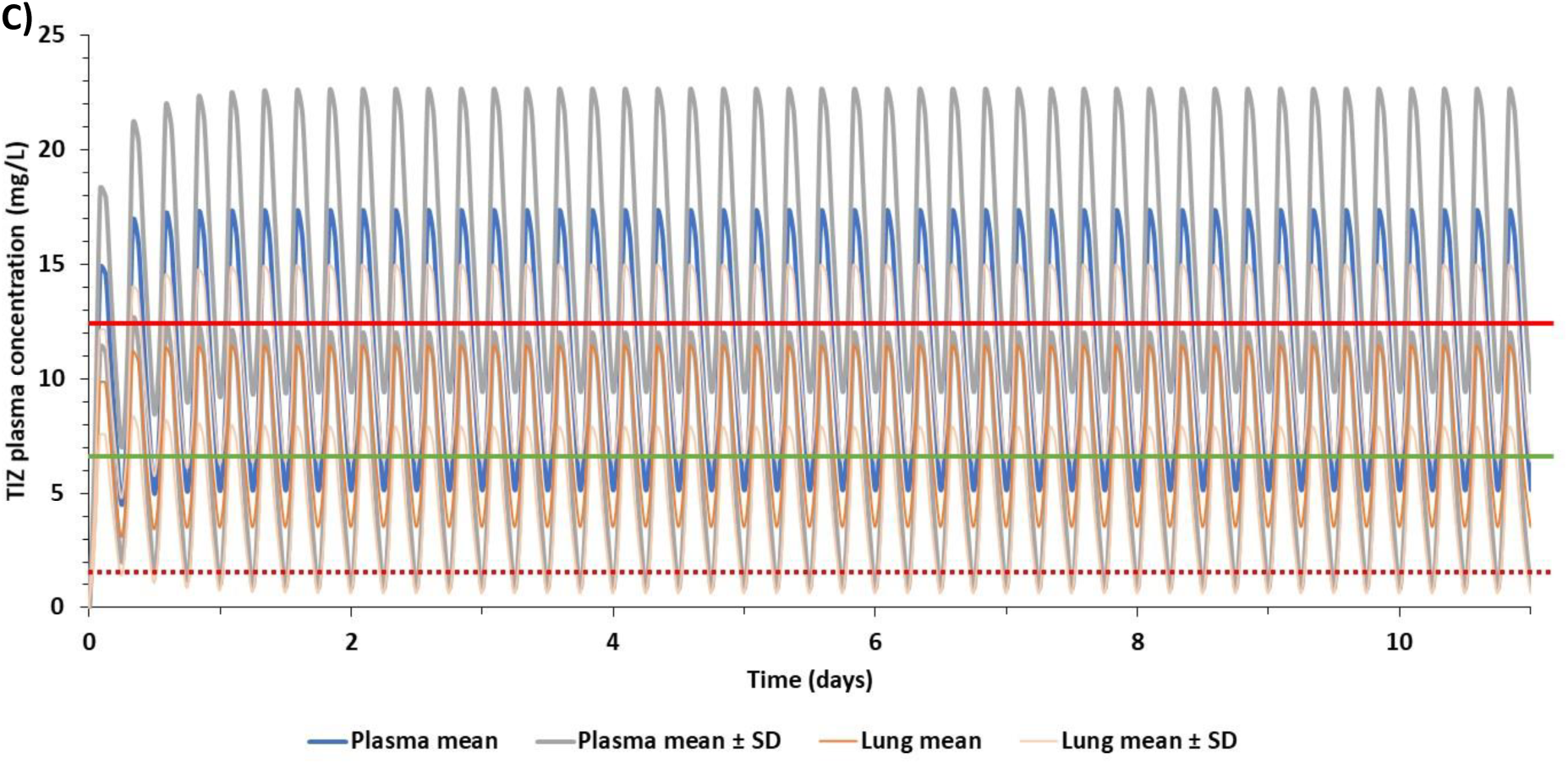
Predicted plasma and lung concentrations for optimal doses during the fed state at different regimens reaching steady state – A) 2900 mg BID, B) 1600 mg TID and C) 1200 mg QID. TIZ – tizoxanide, SD - standard deviation, solid red line indicates clinical C_max_ of 1 g single dose at fested state [29], solid green line represents clinical C_max_ of 500 mg single dose [35] at fested state and the dotted red line represents the EC_90_ of nitazoxanide for SARS-CoV-2 [50].

